# Acute gastrointestinal injury in critically ill patients with coronavirus disease 2019 in Wuhan, China

**DOI:** 10.1101/2020.03.25.20043570

**Authors:** Jia-Kui Sun, Lei Zou, Ying Liu, Wen-Hao Zhang, Jing-Jing Li, Xiao-Hua Kan, Jiu-Dong Chen, Qian-Kun Shi, Shou-Tao Yuan, Wei Gu, Jian-Wei Qi

**Affiliations:** Department of Intensive Care Unit, Nanjing First Hospital, Nanjing Medical University, Nanjing, Jiangsu Province, China; Department of Intensive Care Unit, Lishui People’s Hospital, Nanjing, Jiangsu Province, China; Department of Isolation Units, Tongji Hospital, Huazhong University of Science and Technology, Wuhan, Hubei Province, China

**Keywords:** Gastrointestinal injury, Organ dysfunction, Septic shock, Critically ill, COVID-19

## Abstract

**Background:** To investigate the prevalence and outcomes of acute gastrointestinal injury (AGI) in critically ill patients with coronavirus disease 2019 (COVID-19).

**Methods:** In this clinical retrospective study, demographic data, laboratory parameters, AGI grades, clinical severity and outcomes were collected. The primary endpoints were AGI incidence and 28-day mortality, the secondary endpoints were organ dysfunction and septic shock incidence.

**Results:** From February 10 to March 10 2020, 83 critically ill patients of 1314 patients with COVID-19 were enrolled. Seventy-two (86.7%) patients had AGI during hospital stay, of them, 30 had AGI grade I, 35 had AGI grade II, 5 had AGI grade III, and 2 had AGI grade IV. The incidence of AGI grade II and above was 50.6%. As of March 16, 40 (48.2%) patients died within 28 days of admission, the median hospital stay was 12.0 days, ranging from 3 days to 27 days. Multiple organ dysfunction syndrome developed in 58 (69.9%) patients, septic shock in 16 (19.3%) patients. Patients with worse AGI grades had worse clinical variables, higher septic shock incidence and 28-day mortality. Sequential organ failure assessment scores (SOFA) (95% CI, 1.374-2.860; *P* <0.001), white blood cell (WBC) counts (95% CI, 1.037-1.379; *P* =0.014), duration of mechanical ventilation (MV) (95% CI, 1.020-1.340; *P* =0.025) were risk factors for the development of AGI grade II and above. Non-survivors were accompanied by higher incidence of AGI grade III to IV than survivors (17.5% vs. 0.0%, *P* =0.004).

**Conclusions:** The AGI incidence was 86.7%, and hospital mortality was 48.2% in critically ill patients with COVID-19. SOFA scores, WBC counts, and duration of MV were risk factors for the development of AGI grade II and above. Patients with worse AGI grades had worse clinical severity variables, higher septic shock incidence and 28-day mortality.

## Introduction

In December 2019, clusters of acute pneumonia cases of unclear etiology were identified in Wuhan, the capital of Hubei province in China(1-3). The pathogen was reported as a novel coronavirus that was named severe acute respiratory syndrome coronavirus 2 (SARS-CoV-2). The World Health Organization (WHO) has made the assessment that coronavirus disease 2019 (COVID-19) can be characterized as a pandemic because the disease is spreading rapidly around the world (4). As of March 02, a total of 80151 cases (2943 deaths) were confirmed in China, including 49426 cases (2251 deaths) in Wuhan (5).

The National Health Commission of China has issued a series of diagnosis and treatment recommendations and suggested classifying the disease into four grades: mild, moderate, severe and critical(5). Recent studies have reported the clinical characteristics and prognosis of COVID-19 with varied severity(1, 2, 6-8). Most of the critically ill patients had organs injury, including acute respiratory distress syndrome (ARDS), acute kidney injury (AKI), cardiac injury, or liver dysfunction(9). During our clinical work against the epidemic of COVID-19 in Wuhan, we observed that numerous patients had gastrointestinal symptoms during the course of disease development. It is known that gastrointestinal dysfunction is closely related to adverse outcomes in common critically ill patients. However, few literatures on acute gastrointestinal injury (AGI) were reported in critically ill patients with COVID-19. In this study, we investigated the prevalence and outcomes of AGI in critically ill patients with COVID-19 who were admitted to Guanggu District of Wuhan Tongji Hospital.

## Methods

### Patients

From February 10 to March 10 2020, adult patients (age ≥18 years) with confirmed COVID-19 admitted to our specialized isolation units and intensive care unit (ICU), Guanggu District of Wuhan Tongji hospital were enrolled in this clinical retrospective study. Patients with chronic organ dysfunction (e.g., hepatic or renal dysfunction), immunodeficiency, terminal cancer, and patients with a history of long-term use of hormones were excluded. The written informed consent was waived by our institutional review board because this was a retrospective study for emerging infectious disease.. The diagnosis of COVID-19 was according to WHO interim guidance and recommendations of National Health Commission of China(4, 5), and identified by RNA detection of the SARS-CoV-2 in clinical laboratory of Tongji hospital. The clinical outcomes (e.g., mortality, AGI incidence) were monitored up to March 16, 2020.

### Definitions

A identified case of COVID-19 was defined as a positive finding on real-time reverse-transcriptase–polymerase-chain-reaction (RT-PCR) assay of nasal and pharyngeal swab specimens(4, 5, 7). Only laboratory-confirmed cases were enrolled in the analysis. The clinical classification of critical COVID-19 was in accordance with the Chinese recommendations(5): Meeting any of the following, I, respiratory failure with mechanical ventilation (MV); II, shock; III, multiple organs failure requiring ICU treatment. AGI was defined as a malfunction of the gastrointestinal tract due to acute illness and was categorized into four grades according to its severity(10). This AGI grading system was based mainly on gastrointestinal symptoms, intra-abdominal pressure, feeding tolerance or not. AGI grade I was defined as an increased risk of developing gastrointestinal dysfunction or failure (a self-limiting condition); AGI grade II was defined as gastrointestinal dysfunction (a condition that requires interventions); AGI grade III was defined as gastrointestinal failure (GI function cannot be restored with interventions); AGI grade IV was defined as dramatically manifesting gastrointestinal failure (a condition that is immediately life-threatening)(10). Sepsis was defined as life-threatening organ dysfunction caused by a dysregulated host response to infection, septic shock was defined as a subset of sepsis with circulatory and cellular/metabolic dysfunction associated with a higher risk of mortality(11). The diagnostic criteria of ARDS were in accordance with the Berlin definitions(12). The definitions of AKI were based on the 2012 Kidney Disease: Improving Global Outcomes guidelines(13). Cardiac injury was defined if serum levels of cardiac biomarkers (e.g., troponin I) were above the 99th percentile reference upper limit or new abnormalities were found in electrocardiography and echocardiography(2). Liver injury was defined if serum levels of hepatic biomarkers (e.g., alanine aminotransferase) were above than twice of the reference upper limit or disproportionate elevation of alanine aminotransferase and aspartate aminotransferase levels compared with alkaline phosphatase levels(14). Multiple organ dysfunction syndrome (MODS) was defined as the combined dysfunction of two or more organs.

### Data collection

The baseline clinical characteristics, including sex, age, days from onset to admission, initial symptoms or signs, body mass index (BMI) were collected from electronic medical and nursing records, and all laboratory tests were performed according to the clinical needs of patients. The acute physiology and chronic health evaluation (APACHE) II score, sequential organ failure assessment (SOFA) score, serum levels of C-reactive protein (CRP), D-dimer, white blood cells (WBC) count, lymphocyte count, procalcitonin (PCT), and blood lactate within 24 hours of admission were recorded. The RT-PCR assay of viral RNA was performed by a commercial kit (Tianlong, Xi’an, China) according to the manufacturer’s instructions. All the laboratory parameters were detected by clinical laboratory of Tongji hospital. Moreover, The numbers of patients with AGI (grades), ARDS, AKI, cardiac injury, liver injury, septic shock, MODS, and patients receiving MV or continuous renal replacement therapy (CRRT) during hospital stay were also recorded. The primary endpoints were AGI incidence and 28-day mortality. The secondary endpoints were MODS and septic shock incidence.

### Statistical analysis

The Kolmogorov-Smirnov test was first performed to test the normal distribution of the data. Normally distributed data were expressed as the means ± standard deviation and were compared by t tests. Abnormally distributed data were expressed as the medians (interquartile ranges, IQR) and were compared by the Mann-Whitney U test or the Kruskal-Wallis test. Categorical variables were presented as absolute numbers or percentages and were analyzed using the χ^2^ test or Fisher’s exact test. To take into account the repeated nature of the variables, analysis of variance (ANOVA) for repeated measurements of the general linear model was implemented. Pearson test was used to analyze the correlation between two variables. To determine the risk factors associated with AGI grade II and above, we performed a series of several univariate logistic regression analyses using the above-mentioned variables. Variables with *P* <0.1 in univariate analyses were tested in further multivariate logistic regression analyses. Receiver operating characteristic (ROC) curves were used to evaluate the associations between AGI and MODS, septic shock, and 28-day mortality. Survival curves for up to 28 days after admission and 60 days from disease onset were generated using the Kaplan–Meier method and were compared by the log-rank test. IBM Statistical Package for the Social Sciences (SPSS, version 22.0, NY, USA) software was used for statistical analysis, and *P* <0.05 was considered statistically significant. The statistical methods of this study were reviewed by Qiao Liu, a biostatistician from the Center for Disease Control and Prevention of Jiangsu Province in China.

## Results

As shown in Figure 1, a total of 83 critically ill patients with confirmed COVID-19 were enrolled in this clinical retrospective study. The median age was 70 (IQR, 60-79) years, and most patients were male 59 (71.1%). Fever (33/83, 39.8%) and cough (18/83, 21.7%) were the main initial symptoms. Seventy-two (86.7%) patients had AGI during hospital stay, of them, 30 had AGI grade I, 35 had AGI grade II, 5 had AGI grade III, 2 had AGI grade IV. The incidence of AGI grade II and above was 50.6% (42/83). The detailed clinical data of the patients were presented in Table 1. As of March 16, 40 (48.2%) patients died within 28 days of admission, the median hospital stay of them was 12.0 (IQR, 8.0-17.8) days, ranging from 3 days to 27 days. The median duration from disease onset to death was 22.0 (IQR, 15.3-33.0) days, ranging from 8 days to 44 days. ARDS developed in the most patients (77/83, 92.8%), and 5 patients received extracorporeal membrane oxygenation. MODS developed in 58 (69.9%) patients, septic shock in 16 (19.3%) patients.

**Table 1.** Demographic data and clinical parameters.

| Variables | Values |
| --- | --- |
| Age (years) | 70 (60-79) |
| Sex (Male: Female) | 59: 24 |
| Days from onset to admission | 10 (7-15) |
| BMI (kg/m<sup>2</sup>) | 23.8 (22.1-25.1) |
| APACHEII scores | 14.0 (12.0-17.0) |
| SOFA scores | 6.0 (4.0-7.0) |
| Initial symptoms or signs (*n*, %) |  |
| Fever | 33 (39.8%) |
| Cough | 18 (21.7%) |
| Chest tightness or pain | 8 (9.6%) |
| Dyspnea | 7 (8.4%) |
| Fatigue | 6 (7.2%) |
| Diarrhea | 3 (3.6%) |
| Nausea or vomiting | 3 (3.6%) |
| Pharyngalgia/ Myalgia | 3 (3.6%) |
| Abdominal pain | 2 (2.4%) |
| Laboratory parameters |  |
| CRP (mg/L) | 85.9 (49.9-136.0) |
| WBC (10<sup>9</sup>/L) | 10.30 (7.44-14.0) |
| Lymphocyte (10<sup>9</sup>/L) | 0.60 (0.43-0.83) |
| PCT (ng/mL) | 0.27 (0.14-0.50) |
| D-dimer (ug/mL) | 8.29 (2.00-14.19) |
| Lactate (mmol/L) | 2.22 (1.68-2.73) |
| Organs injury (*n*, %) |  |
| ARDS | 77 (92.8%) |
| Liver injury | 15 (18.1%) |
| AKI | 30 (36.1%) |
| Cardiac injury | 37 (44.6%) |
| MODS (*n*, %) | 58 (69.9%) |
| Septic shock (*n*, %) | 16 (19.3%) |
| Duration of MV (days) | 9.0 (6.0-13.0) |
| Duration of CRRT (days) | 0.0 (0.0-5.0) |
| Hospital stay (days) | 18.0 (11.0-29.0) |
| Death (*n*, %) | 40 (48.2%) |

BMI, body mass index; APACHEII, acute physiology and chronic health evaluation II; SOFA, sequential organ failure assessment; CRP, C-reactive protein; WBC, white blood cell; PCT, procalcitonin; ARDS, acute respiratory distress syndrome; AKI, acute kidney injury; MV, mechanical ventilation; CRRT, continuous renal replacement therapy.

**Figure 1.**
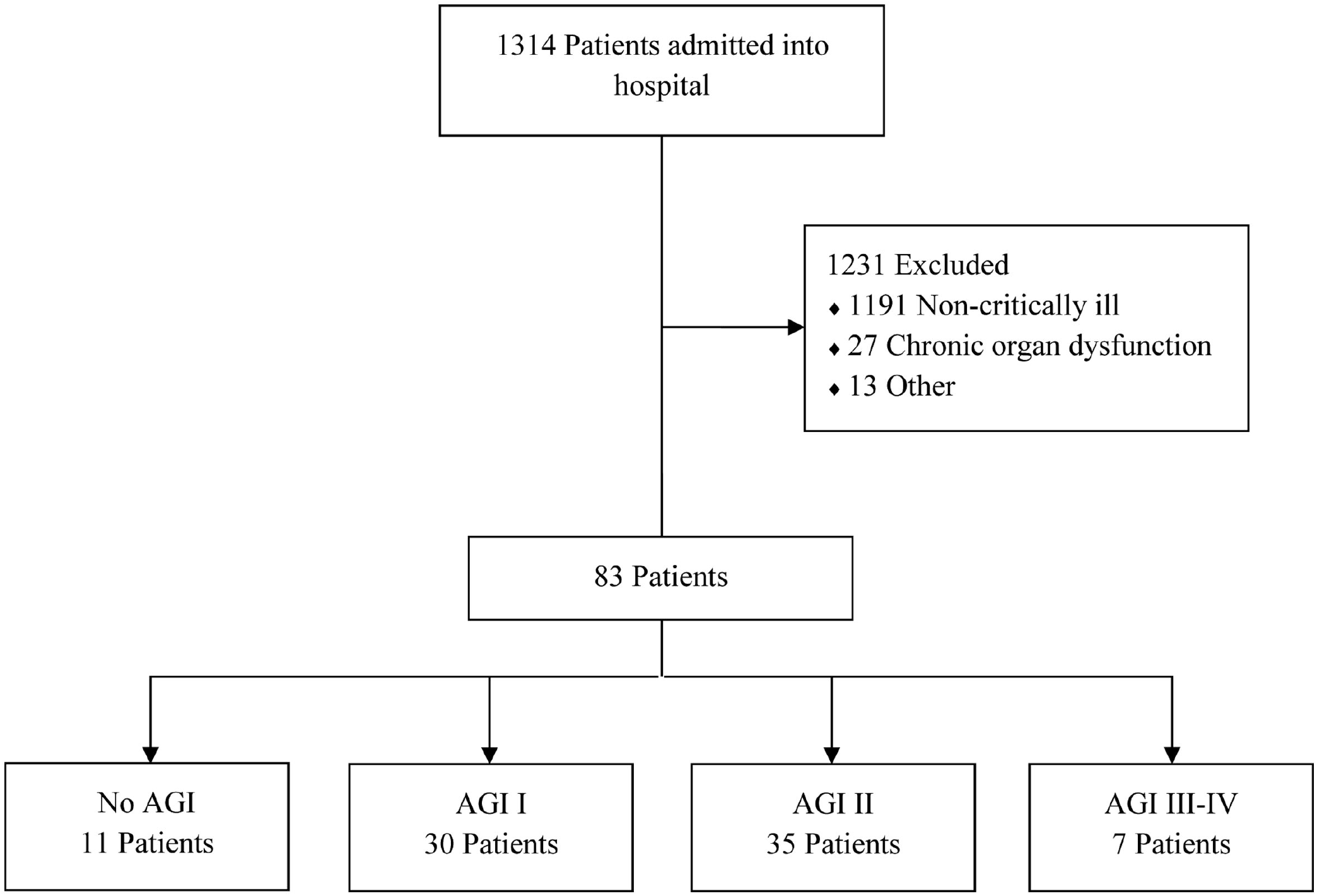
The flow diagram of participants..

### AGI grades and clinical variables

We divided patients into four groups based on the AGI grades: No AGI (*n* =11), AGI grade I (*n* =30), AGI grade II (*n* =35), and AGI grade III to IV (*n* =7). As shown in Table 2, significant differences in APACHEII scores, SOFA scores, WBC counts, and D-dimer levels were found among the four groups (*P* <0.05). Statistical differences in CRP (*P* =0.024) and PCT (*P* =0.033) were only found between group AGI grade I and grade III to IV. Significant differences in lactate levels were found between group no AGI and AGI grade II (*P* =0.027) or grade III to IV (*P* =0.009). Statistical differences in lymphocyte counts were found between group no AGI and AGI grade I (*P* =0.028) or grade II (*P* =0.007). No differences in BMI were found among the four groups (*P* >0.05).

**Table 2.** The AGI grades and clinical variables.

| Variables | No AGI<br>(n =11) | AGI I<br>(n =30) | AGI II<br>(n =35) | AGI III-IV<br>(n =7) | P value |
| --- | --- | --- | --- | --- | --- |
| BMI (kg/m<sup>2</sup>) | 23.9 (22.5-24.9) | 23.8 (21.5-26.1) | 23.8 (22.1-24.9) | 22.4 (21.6-23.1) | 0.426 |
| APACHEII scores | 11.0 (9.0-12.0) | 13.0 (10.0-14.3) | 15.0 (13.0-18.0) | 18.0 (16.0-21.0) | <0.001 |
| SOFA scores | 3.0 (3.0-4.0) | 5.0 (4.0-6.0) | 6.0 (5.0-9.0) | 10.0 (7.0-12.0) | <0.001 |
| CRP (mg/L) | 82.7 (46.1-136.0) | 72.3 (47.3-113.2) | 99.8 (54.6-145.2) | 130.4 (87.3-221.0) | 0.155 |
| WBC (10<sup>9</sup>/L) | 7.9 (6.4-11.3) | 8.7 (7.2-11.8) | 11.9 (8.1-15.1) | 16.6 (12.9-25.5) | 0.001 |
| Lymphocyte (10<sup>9</sup>/L) | 0.85 (0.62-1.12) | 0.66 (0.43-0.82) | 0.51 (0.38-0.76) | 0.59 (0.37-0.85) | 0.044 |
| PCT (ng/mL) | 0.17 (0.08-0.49) | 0.19 (0.13-0.41) | 0.29 (0.16-0.50) | 0.41 (0.31-0.97) | 0.079 |
| D-dimer (ug/mL) | 2.65 (0.97-7.06) | 4.60 (1.72-12.30) | 11.90 (4.64-22.00) | 15.51 (3.76-22.00) | 0.001 |
| Lactate (mmol/L) | 1.17 (1.09-1.88) | 2.09 (1.68-2.44) | 2.25 (1.94-2.99) | 2.55 (1.92-3.84) | 0.038 |
| MV days | 6.0 (4.0-11.0) | 7.0 (5.8-10.5) | 10.0 (6.0-14.0) | 14.0 (11.0-19.0) | 0.029 |
| CRRT days | 0.0 (0.0-4.0) | 0.0 (0.0-3.0) | 0.0 (0.0-7.0) | 6.0 (5.0-8.0) | 0.045 |
| Days from onset to admission | 10.0 (7.0-15.0) | 10.0 (6.0-14.3) | 10.0 (7.0-15.0) | 15.0 (10.0-21.0) | 0.263 |
| Hospital stay (days) | 30.0 (28.0-34.0) | 13.5 (9.0-24.0) | 18.0 (11.0-31.0) | 19.0 (13.0-28.0) | 0.020 |
| MODS(n, %) | 6 (54.5%) | 18 (60.0%) | 27 (77.1%) | 7 (100.0%) | 0.089 |
| Septic shock (n, %) | 0 (0.0%) | 2 (6.7%) | 9 (25.7%) | 5 (71.4%) | 0.001 |
| Death (n, %) | 0 (0.0%) | 14 (46.7%) | 19 (54.3%) | 7 (100.0%) | 0.037 |

AGI, acute gastrointestinal injury; BMI, body mass index; APACHEII, acute physiology and chronic health evaluation II; SOFA, sequential organ failure assessment; CRP, C-reactive protein; WBC, white blood cell; PCT, procalcitonin; ARDS, acute respiratory distress syndrome; AKI, acute kidney injury; MV, mechanical ventilation; CRRT, continuous renal replacement therapy; MODS, multiple organ dysfunction syndrome.

Patients without AGI had longer hospital stay than those with AGI grade I (*P* =0.002), II (*P* =0.022), and III to IV (*P* =0.012). Patients with AGI grade III to IV had longer days of MV and CRRT than those without AGI (*P* =0.011, 0.013) and with AGI grade I (*P* =0.009, 0.007). No differences in days from onset to admission were found among the four groups (*P* >0.05).

Correlation analysis showed that the AGI grades were positively correlated with MV days (R =0.377, *P* <0.001), APACHEII (R =0.590, *P* <0.001) and SOFA scores (R =0.662, *P* <0.001), WBC counts (R =0.433, *P* <0.001), CRP (R =0.261, *P* =0.017) and D-dimer levels (R =0.425, *P* <0.001).

### AGI grades and clinical outcomes

As shown in Table 2, patients with AGI grade III to IV had higher septic shock incidence than those without AGI (*P* =0.002) and with AGI grade I (*P* =0.001) and II (*P* =0.031). Significant differences in 28-day mortality were found among the four groups (*P* <0.05) except for group AGI grade I and II (*P* =0.540). No differences in MODS incidence were found among the four groups (*P* >0.05). Non-survivors were accompanied by higher incidence of AGI grade III to IV than survivors (17.5% vs. 0.0%, *P* =0.004) (Table 3). Whereas survivors had higher incidence of no AGI than non-survivors (25.6% vs. 0.0%, *P* <0.001) (Table 3).

**Table 3.** The incidences of different AGI grades in non-survivors and survivors.

| AGI grades | Non-survivors<br>(n =40) | Survivors<br>(n =43) | P value |
| --- | --- | --- | --- |
| No AGI | 0 (0.0%) | 11 (25.6%) | <0.001 |
| AGI I | 14 (35.0%) | 16 (37.2%) | 0.834 |
| AGI II | 19 (47.5%) | 16 (37.2%) | 0.343 |
| AGI III-IV | 7 (17.5%) | 0 (0.0%) | 0.004 |

AGI, acute gastrointestinal injury.

To determine the risk factors associated with AGI grade II and above, univariate logistic regression was performed using the above-mentioned variables (sex, age, days from onset to admission, BMI, APACHEII scores, SOFA scores, CRP, D-dimer, WBC counts, lymphocyte counts, PCT, blood lactate, MV days, CRRT days, and hospital stay). Variables with *P* <0.1 in univariate analyses were tested in further multivariate logistic regression analyses. As shown in Table 4, three variables (SOFA scores, WBC counts, MV days) were established as independent risk factors for the development of AGI grade II and above.

**Table 4.** Independent factors associated with AGI grade II and above in multivariate logistic regression analysis

| Variables | OR | 95% CI | P value |
| --- | --- | --- | --- |
| SOFA scores | 1.982 | 1.374-2.860 | <0.001 |
| WBC counts | 1.196 | 1.037-1.379 | 0.014 |
| MV days | 1.169 | 1.020-1.340 | 0.025 |

AGI, acute gastrointestinal injury; SOFA, sequential organ failure assessment; WBC, white blood cell; MV, mechanical ventilation.

The ROC curves were performed to evaluate the associations between AGI and clinical outcome variables. The area under curves (AUCs) of MODS, septic shock, and 28-day mortality were 0.659 (*P* =0.022), 0.793 (*P* <0.001), and 0.716 (*P* =0.001), respectively (Figure 2). Significant differences in 28-day mortality after admission (*P* =0.002) and 60-day mortality after disease onset (*P* =0.003) were found between group no AGI (*n* =11) and AGI (*n* =72). As shown in Figure 3, statistical differences in the 28-day mortality (*P* =0.037) and the 60-day mortality (*P* =0.049) were also found between group AGI grade I to below (*n* =41) and AGI grade II to IV (*n* =42).

**Figure 2.**
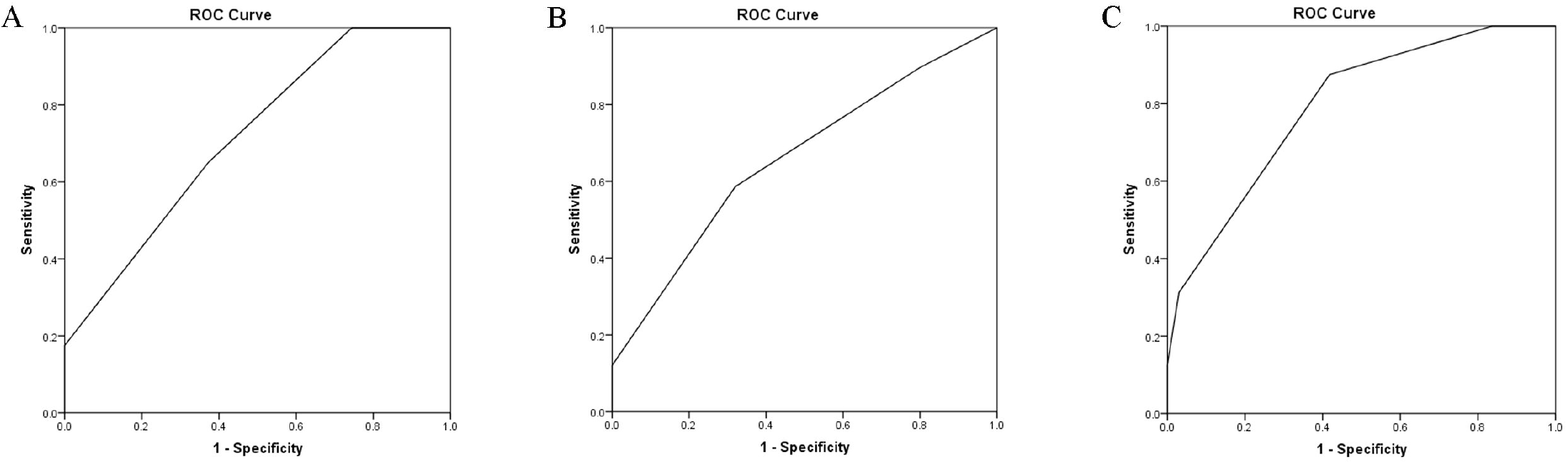
The areas under the receiver operating characteristic curves of multiple organ dysfunction syndrome (Fig 3A), septic shock (Fig 3B), and 28-day mortality (Fig 3C) were 0.659 (*P* =0.022), 0.793 (*P* <0.001), and 0.716 (*P* =0.001), respectively.

**Figure 3.**
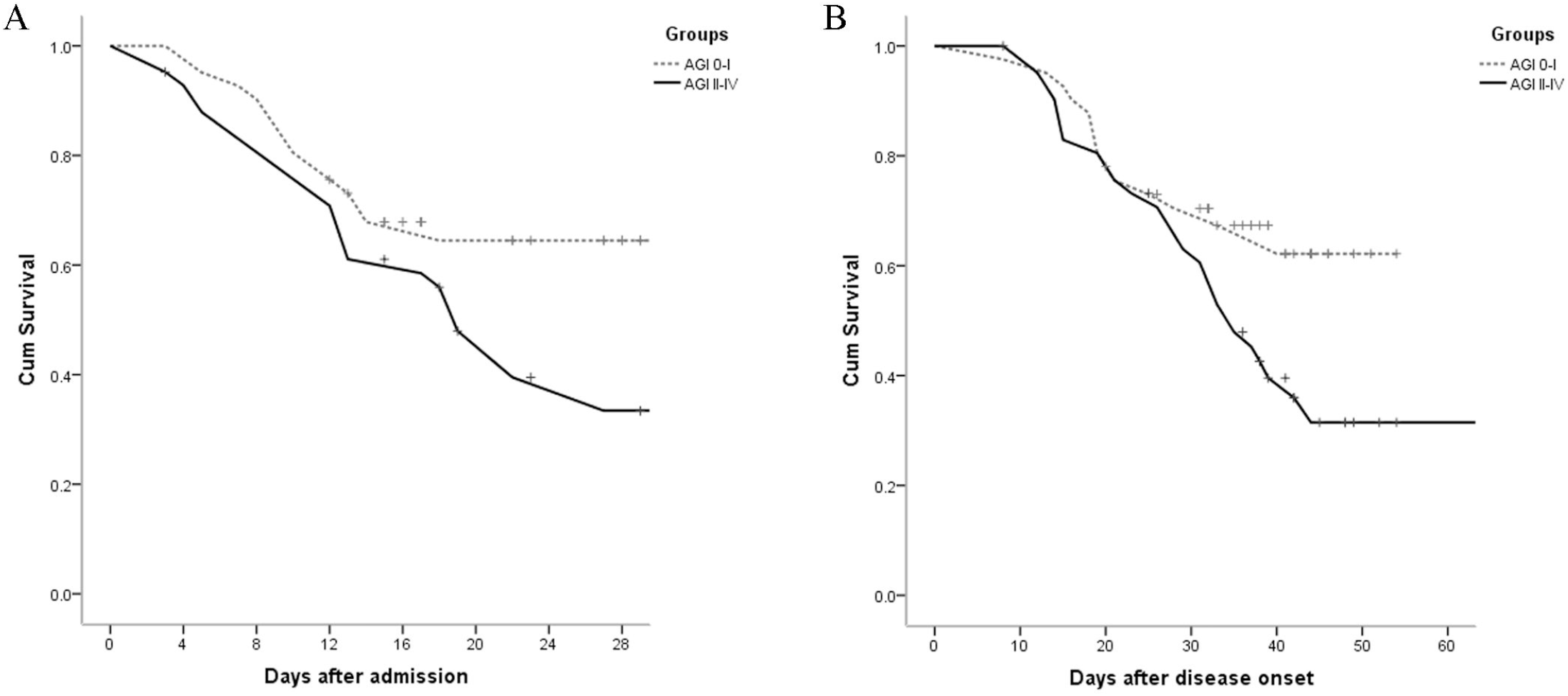
Significant differences in 28-day mortality after admission (*P* =0.037) and 60-day mortality after disease onset (*P* =0.049) were found between group AGI grade I to below (*n* =41) and AGI grade II to IV (*n* =42).

## Discussion

This clinical retrospective study investigated the prevalence and outcomes of AGI in critically ill patients with COVID-19. 86.7% of the patients had AGI, and 50.6% had AGI grade II and above during hospital stay. We found that patients with worse AGI grades had worse clinical severity variables, higher septic shock incidence, higher 28-day mortality after admission and 60-day mortality after disease onset. SOFA scores, WBC counts, duration of MV were risk factors for the development of AGI grade II and above. The 28-day mortality, MODS incidence, and septic shock incidence of critically ill patients were 48.2%, 69.9%, and 19.3%, respectively. Non-survivors were accompanied by higher incidence of AGI grade III to IV than survivors.

Most of the critically ill patients with COVID-19 had organs injury, including ARDS and AKI(9). However, few literatures on gastrointestinal injury were reported in patients with COVID-19. Gastrointestinal dysfunction is common and closely related to adverse outcomes in general critically ill patients(10, 15-17). In 2012, the Working Group on Abdominal Problems (WGAP) of the European Society of Intensive Care Medicine developed the definitions and a grading system for AGI in intensive care patients(10). This expert opinion-based AGI grading system had been proven to be a predictor of all-cause mortality (16). To our knowledge, this is the first study to investigate AGI in critically ill patients infected by SARS-CoV-2. Our results showed that AGI incidence was very high in critically ill patients with COVID-19. AGI was also correlated with clinical severity and outcomes of this novel disease. A recent meta-analysis showed that the incidence of AGI was about 40% and the mortality was 33% in critically ill patients(15). The corresponding data of this study were higher than those of the previous reports. This phenomenon indicated that SARS-CoV-2 was more virulent than SARS-CoV, and gastrointestinal tract was also a vital target organ of the virus. However, the underlying mechanisms of SARS-CoV-2 causing organs dysfunction were unknown yet.

Gastrointestinal injury was often caused by an inflammatory reaction, infection or sepsis, severe trauma, shock, pancreatitis, and other critical diseases(10, 15, 16). As the receptor for SARS-CoV, angiotensin-converting enzyme 2 (ACE2) was suggested to be also the receptor for SARS-CoV-2(18). ACE2 was expressed in endothelial cells and smooth muscle cells of almost all organs, especially in lung alveolar cells(18). That’s why the COVID-19 patients were susceptible to ARDS and even MODS. Our findings also showed that ARDS incidence was very high (92.8%), and AGI grades were significantly positive correlated with MV days. Liang et al. reported that ACE2 was highly expressed in small intestine, especially in proximal and distal enterocytes(19). ACE2 expression in the epithelial cells was required for maintaining antimicrobial peptide expression, amino acid homeostasis, and the ecology of gut microbiome in intestine(20). Therefore, gastrointestinal symptoms were also reported in previous studies on COVID-19(7, 8). We believed that these gastrointestinal symptoms were the early manifestations of AGI and should be taken seriously in clinical treatment.

In this study, we found that AGI grades were correlated with APACHEII and SOFA scores, WBC counts, CRP and D-dimer levels. Moreover, SOFA scores, WBC counts, and duration of MV were risk factors for the development of AGI grade II and above. These results indicated that patients with worse AGI grades had more serious virus infection and severe inflammatory response, which may lead to a vicious circle between systemic infection and intestinal barrier damage. D-dimer, a fibrin degradation product, was also considered to be associated with adverse outcomes of COVID-19(21). The abnormal elevation of D-dimer indicated microcirculation disturbance, including microthrombotic formation of intestinal mucosa(21). During our clinical work against the epidemic of COVID-19 in Wuhan, we observed that gastrointestinal hemorrhage developed in several severe patients. We speculated that stress ulcer and intestinal microcirculation disturbance may be causes of the disorder.

Yang et al.(9) reported that ARDS developed in 67%, AKI in 29%, cardiac injury in 23%, and liver dysfunction in 29% of critically ill patients with SARS-CoV-2 pneumonia. Zhou’s study (21) presented that septic shock developed in 20%, ARDS in 31%, AKI in 15%, and cardiac injury in 17% of total patients with COVID-19. Our results showed that ARDS developed in 92.8%, AKI in 36.1%, cardiac injury in 44.6%, and liver injury in 18.1% of the critically ill patients with COVID-19. The incidences of organ injury in this study were higher than those of previous studies, which may suggest that patients with AGI had worse clinical outcomes. The high MODS incidence (69.9%) and hospital mortality (48.2%) of critically ill patients in this study also confirmed this conclusion. Moreover, we found that the hospital days of patients without AGI were significantly longer than that of patients with AGI. It could be explained by the high 28-d mortality in patients with AGI, because the median hospital stay of non-survivors was only 12.0 (IQR, 8.0-17.8) days, ranging from 3 days to 27 days.

Some limitations of the study should be discussed. Because of our single-center retrospective design and small sample size, the results might be inconclusive, and the accuracy should be confirmed by large-scale clinical prospective studies. Moreover, because the study was not based on pathophysiological models, the results were hypothesis generating, the exact mechanisms of AGI in COVID-19 should be tested by more basic experiments. In addition, patients were sometimes transferred late in their illness to our hospital. Lack of effective antivirals and inadequate adherence to standard supportive therapy might have contributed to the poor clinical outcomes in some patients.

## Conclusions

To our knowledge, this is the first study to investigate AGI in critically ill patients with COVID-19. The AGI incidence was 86.7%, and hospital mortality was 48.2% in critically ill patients. SOFA scores, WBC counts, and duration of MV were risk factors for the development of AGI grade II and above. Patients with worse AGI grades had worse clinical severity variables, higher septic shock incidence, and higher hospital mortality.

## Data Availability

The datasets used and/or analyzed during the current study are available from the corresponding author on reasonable request.

## Acknowledgements

The authors thank Qiao Liu for her assistance in the statistical analysis of this study. The authors also thank Li H, Zou J, Dong K, and Jin CC of Tongji hospital for their contributions to this study. In addition, Sun JK and his family especially thank Sun XP for her meticulous care and support during the past ten years.

## Authors’ contributions

Sun JK, Shi QK, Yuan ST, Gu W, and Qi JW designed the research; Sun JK, Zou L, Liu Y, Zhang WH, Li JJ, Kan XH, Chen JD, and Dai L performed the research; Sun JK, Zou L, and Liu Y analyzed the data; Sun JK and Zou L wrote the paper.

## Ethics approval and consent to participate

The study was reviewed and approved by the institutional review board of Nanjing First Hospital and Tongji Hospital, whereas written informed consent was waived because this was a retrospective study.

## Consent for publication

Not applicable.

## References

1. Zhu N, Zhang D, Wang W, et al: China Novel Coronavirus Investigating and Research Team. A Novel Coronavirus from Patients with Pneumonia in China, 2019. N Engl J Med 2020;382:727–733.

2. Wang D, Hu B, Hu C, et al: Clinical Characteristics of 138 Hospitalized Patients With 2019 Novel Coronavirus-Infected Pneumonia in Wuhan, China. JAMA 2020 Feb 7. doi: 10.1001/jama.2020.1585.

3. Li Q, Guan X, Wu P, et al: Early Transmission Dynamics in Wuhan, China, of Novel Coronavirus-Infected Pneumonia. N Engl J Med 2020 Jan 29. doi: 10.1056/NEJMoa2001316.

4. World Health Organization: Coronavirus disease (COVID-19) outbreak (https://www.who.int).

5. National Health Commission of the People’s Republic of China home page (http://www.nhc.gov.cn).

6. Xu XW, Wu XX, Jiang XG, et al: Clinical findings in a group of patients infected with the 2019 novel coronavirus (SARS-Cov-2) outside of Wuhan, China: retrospective case series. BMJ 2020; 368:m606.

7. Guan WJ, Ni ZY, Hu Y, et al: Clinical Characteristics of Coronavirus Disease 2019 in China. N Engl J Med 2020 Feb 28. doi: 10.1056/NEJMoa2002032.

8. Huang C, Wang Y, Li X, et al: Clinical features of patients infected with 2019 novel coronavirus in Wuhan, China. Lancet 2020;395(10223):497–506.

9. Yang X, Yu Y, Xu J, et al: Clinical course and outcomes of critically ill patients with SARS-CoV-2 pneumonia in Wuhan, China: a single-centered, retrospective, observational study. Lancet Respir Med 2020 Feb 24. pii: S2213-2600(20)30079-5.

10. Reintam Blaser A, Malbrain ML, Starkopf J, et al: Gastrointestinal function in intensive care patients: terminology, definitions and management. Recommendations of the ESICM Working Group on Abdominal Problems. Intensive Care Med 2012;38(3):384–394.

11. Rhodes A, Evans LE, Alhazzani W, et al: Surviving Sepsis Campaign: International Guidelines for Management of Sepsis and Septic Shock: 2016. Crit Care Med 2017;45(3):486–552.

12. ARDS Definition Task Force, Ranieri VM, Rubenfeld GD, et al: Acute respiratory distress syndrome: the Berlin Definition. JAMA 2012;307(23):2526–2533.

13. Kidney Disease: Improving Global Outcomes (KDIGO) Acute Kidney Injury Work Group: KDIGO Clinical Practice Guideline for Acute Kidney Injury. Kidney Int Suppl 2012;2:1.

14. Kwo PY, Cohen SM, Lim JK. ACG Clinical Guideline: Evaluation of Abnormal Liver Chemistries. Am J Gastroenterol 2017;112(1):18–35.

15. Zhang D, Li Y, Ding L, et al: Prevalence and outcome of acute gastrointestinal injury in critically ill patients: A systematic review and meta-analysis. Medicine (Baltimore) 2018;97(43):e12970.

16. Hu B, Sun R, Wu A, et al: Severity of acute gastrointestinal injury grade is a predictor of all-cause mortality in critically ill patients: a multicenter, prospective, observational study. Crit Care 2017;21(1):188.

17. Li H, Zhang D, Wang Y, et al: Association between acute gastrointestinal injury grading system and disease severity and prognosis in critically ill patients: A multicenter, prospective, observational study in China. J Crit Care 2016;36:24–28.

18. Chan JF, Kok KH, Zhu Z, et al: Genomic characterization of the 2019 novel human-pathogenic coronavirus isolated from a patient with atypical pneumonia after visiting Wuhan. Emerg Microbes Infect 2020;9(1):221–236.

19. Liang W, Feng Z, Rao S, et al: Diarrhoea may be underestimated: a missing link in 2019 novel coronavirus. Gut 2020 Feb 26. pii: gutjnl-2020-320832.

20. Hashimoto T, Perlot T, Rehman A, et al: ACE2 links amino acid malnutrition to microbial ecology and intestinal inflammation. Nature 2012;487(7408):477–481.

21. Zhou F, Yu T, Du R, et al: Clinical course and risk factors for mortality of adult inpatients with COVID-19 in Wuhan, China: a retrospective cohort study. Lancet 2020 Mar 11. pii: S0140-6736(20)30566-3.

